# Methods utilising routinely collected data for evaluating the effectiveness of population-level public health interventions to improve public awareness of cancer symptoms: protocol for a scoping review

**DOI:** 10.1101/2025.07.24.25332116

**Authors:** Margaret Smith, Sheba Ziyenge, Claire Friedemann Smith, Brian Nicholson, Clare Bankhead

**Author notes:** Corresponding author: Margaret Smith.

## Abstract

**Objective:** The objective of this review is to understand the types of observational study designs and analyses that have been used to investigate the impact of public health interventions that aim to improve early diagnosis of cancer.

**Introduction:** Campaigns to improve public awareness of cancer symptoms need to be evaluated at various stages in the cancer pathway.

**Inclusion criteria:** Analytic observational studies based on databases of routinely collected health data

**Methods:** Abstracts in Ovid Medline and Embase and the Overton policy database from 2010 to the present will be extracted. Full text of potentially relevant sources will be screened. Covidence software will be used to manage the screening and data extraction process. Extracted data will be tabulated and presented graphically. A narrative summary will accompany the tabulated results.

## Introduction

Diagnosing cancer at an earlier stage when it is localised and easier to treat saves lives. It is therefore important for patients to consult with their GP to get possible signs of cancer checked out as soon as possible. Screening programmes are another way in which cancer symptoms can be found early for certain cancers (colon, breast, cervical, and lung). However, lack of awareness or unwillingness to seek help or take up screening due to cancer fear, fatalism, or taboos surrounding the cancer site or testing process means many cancers are identified later than they should be.

Public awareness campaigns, such as the Be Clear on Cancer Campaign aim to increase awareness of cancer symptoms or to increase uptake of screening, either overall or within specific subgroups of the population (1). Evaluation of the success of these campaigns needs to be done at various points in the patient pathway from GP consultations about symptoms and screening attendances through to short and longer-term survival. Randomised controlled designs are the gold standard for inferring causality but they may not be straightforward or economical to conduct. Cluster randomised trials of interventions applied at the population level are one way of controlling for confounding but could be difficult or expensive to follow-up for outcomes that take a long time to develop. Surveys can also be used to evaluate changes in knowledge and intended behaviour changes after cancer awareness campaigns, but they are less useful for evaluating real world impact.

Observational study designs can also be utilised to evaluate campaigns. Simple before-and-after studies are observational designs that can be used to compare outcomes before and after an intervention, but it may be hard to control adequately for pre-existing differences between communities targeted by the intervention and for other factors that may have changed at the same time. Causality cannot necessarily be attributed to the intervention because of this inability to control for both known and unknown confounders and because of the possibility of coincident changes at the time of applying the intervention. Recently methodology for “natural experiments” or “quasi-experimental” non-randomised observational studies that attempt to deal with some of these problems has been increasingly utilised for evaluating public health interventions (2, 3). Interrupted time series, which compare trends pre- and post a sudden intervention (interruption), and difference-in-differences studies, which compare differences between a treatment and control group before and after an intervention, are established methods for evaluating changes in public health policy or public health interventions. There are several papers describing and evaluating methodology for these designs in the context of public health interventions or health policy (4-6). Other approaches that may deal with some of the problems encountered with classical observational studies include propensity score methods, synthetic controls, instrumental variable methods, regression discontinuity designs, and near-far matching (2).

Several routinely collected datasets such as THIN or CPRD Aurum for Primary Care, Hospital Episodes Statistics, Routes to Diagnosis, and ONS mortality data in the UK are now available. Observational studies utilising these large electronic health record databases or other routinely collected data could be a cost-effective way of following up on multiple outcomes after cancer awareness campaigns.

A preliminary search of Ovid Medline for titles containing “systematic review” or “scoping review” and “cancer awareness” and “method*” was conducted and no current or underway systematic reviews or scoping reviews on the topic were identified.

Therefore, we plan to review the observational study designs and statistical approaches that have been used to evaluate public health interventions aimed at improving public awareness of cancer. Our review will focus on evaluations done in pre-existing databases of health records as opposed to prospectively collected datasets such as from surveys or clinical trials.

This protocol was developed using best-practice guidance for developing a scoping review protocol (7, 8) and utilised the JBI template for a scoping review (downloaded from https://jbi.global/scoping-review-network/resources).

## Review question

Which methods are used with routinely collected data to evaluate the effectiveness of population-level public health interventions that aim to improve public awareness of cancer symptoms?

- Population/participants: All
- Concept: Methodological approaches in observational studies for assessing effectiveness.
- Context: Health interventions applied at the population or community level to improve awareness of cancer symptoms or cancer screening.

## Eligibility criteria

### Participants

This a methodological review of study design and analysis so there are no restrictions on participants.

### Concept

Methodological approaches for assessing effectiveness in observational studies that have used routinely collected data.

This scoping review will consider all analytical observational studies for inclusion including quasi-experimental studies, interrupted time series, before-after studies, retrospective cohort studies, and cross-sectional studies. Studies using a mixture of data sources for different outcomes will be included as long as at least one meets the study criteria. Impact can be evaluated over any period of time and at any point in the cancer pathway e.g. GP consultations for symptoms, urgent referrals, screening uptake, cancer survival.

Specific exclusions: evaluation methods that use prospectively collected data e.g. clinical trials or surveys or studies based on questionnaires; studies that only evaluate impact by monitoring social media, for example Google trends analysis; studies that only contain an economic evaluation.

### Context

Health interventions applied at the population or community level to improve early diagnosis of cancer through increasing public awareness of cancer symptoms or of the benefits of screening. Interventions aimed at the general population will be included. For example, health education, social marketing or media campaigns that improve awareness of cancer symptoms and encourage people to consult about symptoms, or campaigns that encourage people to take up opportunities for routine screening.

Specific exclusions: evaluations of screening programmes themselves (as opposed to interventions aimed to promote uptake of screening); cancer prevention interventions e.g. tobacco control or vaccination; interventions that are not directly aimed at increasing public awareness of cancer symptoms or screening programmes e.g. changes in practice guidelines; legislative changes; or interventions that change access to care e.g. changes in taxation or co-payments.

### Types of evidence sources

The review will include studies in the Medline and Embase bibliographic databases and additional studies published in the grey literature. This review will consider analytical observational studies published in the English language. Studies published since 2010 until the present will be included. 2010 predates the Be Clear on Cancer Campaign that ran from 2011 to 2018. Studies using routinely collected data would have been very uncommon prior to 2010 because these databases were not so well known, and the methodology for different observational study designs has developed considerably since 2010. The English language criterion is for feasibility.

Descriptive studies and studies that do not make a specific comparison (e.g. with a control group or pre-intervention) will be excluded. Qualitative studies and text and opinion papers will be excluded. Systematic reviews will also be excluded, but references will be scanned for potential studies to include in this review.

### Summary of eligibility criteria

#### Inclusion criteria

- Studies that evaluate the impact of public health interventions that aim to improve awareness of cancer or cancer symptoms
- Analytical studies:
- Observational studies using routinely collected healthcare data

#### Exclusion criteria

- Not evaluating an intervention or wrong type of intervention or intervention not aimed at the general public
  ∘ Evaluating a screening programme (rather than its promotion), changes in practice guidelines, legislative changes, changes in taxation or co-payments
- Wrong type of study
  ∘ Descriptive study, qualitative study, trial, health economics study
- Studies that don’t use routinely collected healthcare data
  ∘ Studies that only use prospectively collected data e.g. surveys or questionnaires, studies that monitor social media data

## Methods

The proposed scoping review will be conducted in accordance with the JBI methodology for scoping reviews.(8)

### Search strategy

A limited search of Ovid Medline was undertaken to identify articles on the topic. The text words contained in the titles and abstracts of relevant articles, the index terms used to describe the articles, supplemented by terms added based on reviewer expertise were used to develop a full search strategy for OVID Medline. These were supplemented by some relevant MESH terms found in articles on the topic. The search strategy includes four sets of search terms:

1. Cancer
2. The intervention e.g. cancer awareness campaign
3. Terms indicating evaluation e.g. “impact”
4. The outcome that was evaluated e.g. primary care consultations

Searches 1) and 2) cover the “Context” part of the PCC. Searches 3) and 4) cover the “Concept”. There are no restrictions on population.

Searches 1) to 4) were then combined with “AND”. Further search terms were added to refine the search to remove studies that were about randomised controlled trials or surveys or questionnaires. Limits on date and language were applied. The draft search strategy for Ovid Medline is given in Appendix 1. The search strategy, including all identified keywords and index terms, will be adapted for Ovid Embase. We will also search the Overton database for additional relevant reports.

### Study/Source of evidence selection

Following the search, all identified citations will be collated and uploaded into Covidence systematic review software and duplicates removed (9). Titles and abstracts will then be screened by two independent reviewers for assessment against the review inclusion criteria. Potentially relevant sources will be retrieved in full, and their citation details imported into the Covidence software. The full text of selected citations will be assessed in detail against the inclusion criteria by two independent reviewers. Reasons for exclusion of sources of evidence at full text that do not meet the inclusion criteria will be recorded and reported in the scoping review.

Pilot tests at each selection stage will be used to clarify the eligibility criteria and definitions/elaboration document that will be used by the entire team. Any disagreements that arise between the reviewers at each stage of the selection process will be resolved through discussion, or with an additional reviewer and the definitions/elaboration document will be modified as necessary. The results of the search and the study inclusion process will be reported in full in the final scoping review and presented in a PRISMA flow diagram (10).

### Data extraction

Data will be extracted from the selected studies by two independent reviewers using a data extraction tool developed by the reviewers. The tool will be piloted by two reviewers on a random sample of studies before release to the rest of the team. The data extraction tool will be modified and revised as necessary during the process of extracting data from each included evidence source. Modifications will be detailed in the scoping review. Any disagreements that arise between the reviewers will be resolved through discussion, or with an additional reviewer/s.

#### Data extracted will include the following

- Bibliographic details
- Study design e.g. pre-post, interrupted time series
- Details of database used e.g. CPRD, THIN
- Details of the study (population, intervention(s), control group(s), outcome(s))
- Details of study size and number of data points
- Time period of data
- Type of outcome e.g. continuous, binary, count, rates, time to event
- Statistical analysis methods used
- Methods to control for confounding
- Additional features of design and analysis following good practice guidelines (if these exist).

## Data Availability

This is a protocol for a scoping review so there are no data.

## Data analysis and presentation

Selected studies will be grouped by the study design used. Extracted data will be tabulated and presented graphically. A narrative summary will accompany the tabulated and/or charted results and will describe how the results relate to the reviews objective and question/s.

## Deviations from the protocol

Revisions to the registered protocol will be fully recorded and published online along with the original protocol.

## Acknowledgements

Nia Roberts, Bodleian Health care Libraries, University of Oxford gave advice on developing the searches.

## Author contributions

CB and BN conceived the study. MS led in developing the concept, designing the study, and developing the searches, and wrote the first draft of this manuscript. SZ contributed to developing the concept, design of the study, and the searches. CFS advised on developing the searches. All authors commented on earlier drafts of the protocol. All authors saw and approved the final version.

