## Appendix 1 for "Methods utilising routinely collected data for evaluating the effectiveness of population-level public health interventions to improve public awareness of cancer symptoms: protocol for a scoping review"

### Appendices

#### Appendix I: Search strategy

##### Medline (Ovid MEDLINE® Epub Ahead of Print, In-Process & Other Non-Indexed Citations, Ovid MEDLINE® Daily and Ovid MEDLINE®) 1946 to present

| | | N records retrieved |
|---|---|---|
| 1 | ("35570375" or "35384107" or "34718357" or "34382254" or "31839675" or "30813110" or "29950525" or "29067155" or "26072663" or "25461805" or "23514877" or "19942689" or "26096269" or "22052579" or "26511591" or "29607059" or "35063941" or "22823531").ui. | 18 |
| 2 | exp *Neoplasms/ | 3652247 |
| 3 | (cancer? or carcinoma? or tumo?r? or malignan* or neoplas* or oncol*).ti,kf. | 2891193 |
| 4 | 2 or 3 | 4328639 |
| 5 | (campaign* or promotion? or advertis* or initiative? or strateg* or intervention? or drive or education or broadcast? or social marketing).ti,kf. | 843054 |
| 6 | ((campaign* or promotion? or advertis* or initiative? or strateg* or intervention? or drive or education or broadcast? or social marketing) adj3 ((awareness or knowledge) adj3 (cancer or symptom?))).mp. | 705 |
| 7 | ((campaign* or promotion? or advertis* or initiative? or strateg* or intervention? or drive or education or broadcast? or social marketing) adj3 cancer screening).mp. | 2102 |
| 8 | Mass Media/ or Advertising/ or Social Marketing/ | 29874 |
| 9 | Health Promotion/ | 85225 |
| 10 | ("Detect Cancer Earlier" or "Find Cancer Early" or "Be Cancer Aware" or "SunSmart" or "Be Clear on Cancer" or "Help Us Help You" or "Be the Early Bird" or "Inside Knowledge" or Movember or "National Colorectal Cancer Awareness Month" or "SunSmart" or "Breast Cancer Awareness Month" or "Screen for Life" or "Slip, Slop, Slap").mp. | 329 |
| 11 | (impact or evaluat* or effectiveness or influence or efficacy or appraisal or assessment).mp. | 9649644 |
| 12 | Program Evaluation/ or Evaluation Study/ | 326666 |
| 13 | ((survival or mortality) adj2 ("one year" or 1-year or 12-month? or twelve month?)).mp. | 33228 |
| 14 | ((refer* adj2 urgent) or 2WW or two week? wait or 2-week? wait).mp. | 1211 |
| 15 | ((refer* or attend* or consult* or present*) adj3 (general practitioner? or physician? or clinician? or primary care or GP?)).mp. | 54694 |
| 16 | (screen* adj5 (uptake or acceptance or adhere* or nonadhere* or complian* or noncomplian* or comply or attend* or nonattend* or participat* or number? or complet*)).mp. | 40830 |
| 17 | (rate? adj2 (death or mortality or incidence or referral or consultation or diagnosis or hospitalisation or uptake or test*)).mp. | 362455 |
| 18 | ((earl* adj3 diag*) or stage shift).mp. | 205068 |
| 19 | (screen* and (kit adj2 return)).mp. | 45 |
| 20 | Early Detection of Cancer/ | 44035 |
| 21 | Health Knowledge, Attitudes, Practice/ | 136642 |
| 22 | ((randomis* or randomiz* or clinical) adj2 trial?).ti,kf. | 318189 |
| 23 | (questionnaire or survey).ti,kf. | 256809 |
| 24 | Clinical Trial/ or Randomized Controlled Trial/ | 963295 |
| 25 | Surveys and Questionnaires/ | 623471 |
| 26 | (4 and (5 or 6 or 7 or 8 or 9 or 10) and (11 or 12) and (13 or 14 or 15 or 16 or 17 or 18 or 19 or 20 or 21)) not (22 or 23 or 24) | 4144 |
| 27 | limit 26 to english language | 3990 |
| 28 | limit 27 to dt=20100101-20250423 | 3371 |
